# Holistic assessment of the effect of alcohol consumption on steatotic liver disease: systematic review and meta-analysis

**DOI:** 10.64898/2026.05.11.26352864

**Authors:** Madina Yerezhepbayeva, Xincheng Li, Jiajing Li, Tianyu Wang, Ibrahim Ayada, Qiuwei Pan

## Abstract

**Background and Aims:** Steatotic liver disease (SLD) is characterized by excessive lipid accumulation in hepatocytes, and alcohol consumption may modify the disease course, but the evidence is inclusive. This systematic review and meta-analysis aimed to holistically evaluate the impact of mild, moderate, and high levels of alcohol consumption on hepatic and extrahepatic outcomes in SLD.

**Methods:** We systematically searched EMBASE, MEDLINE, Web of Science, and the Cochrane Central Register of Controlled Trials for relevant studies. The study outcomes included liver related events, malignancy, mortality and cardiovascular disease among adults with SLD who consumed alcohol.

**Results:** Of 2228 records identified, twenty-six studies comprising 466611 adults with SLD were included. High alcohol consumption was associated with an increased risk of liver-related events compared with abstinence (2.97, 95% CI 1.61–5.50; *p*<0.001), and a similar association was observed among alcohol drinkers overall (HR 1.93, 95% CI 1.60–2.33; *p*<0.001). Moderate alcohol consumption was associated with a higher incidence of malignancy (HR 1.41, 95% CI 1.13–1.78; *p*=0.677). In contrast, mild alcohol consumption was associated with lower all-cause mortality compared with abstinence (HR 0.88, 95% CI 0.78–0.98; *p*=0.001). No association was observed between alcohol consumption and cardiovascular disease incidence or hepatocellular carcinoma

**Conclusions:** Alcohol intake may increase the risk of liver-related complications and cancer risk in individuals with SLD. Mild alcohol consumption was associated with lower all-cause mortality, and alcohol intake showed no association with cardiovascular disease incidence. Further studies are needed to clarify the dose-dependent effects of alcohol on hepatic and extrahepatic outcomes in SLD.

## Introduction

Steatotic liver disease (SLD) represents a spectrum of liver disorders characterized by excessive fat accumulation from various etiologies ^1^. It is highly prevalent, affecting approximately one third of the global adult population ^2^. Recent consensus statements have proposed a reclassification of SLD based on the coexistence of metabolic risk factors and the alcohol consumption ^3^. The term metabolic dysfunction-associated steatotic liver disease (MASLD) has replaced the term non-alcoholic fatty liver disease (NAFLD). MASLD refers to the liver condition characterized by the presence of at least one cardiometabolic risk factor and alcohol consumption below 140 g per week for women and 210 g per week for men. In addition, the revised framework introduced a new category as metabolic dysfunction and alcohol-related liver disease (MetALD), which includes people with SLD who consume alcohol at moderate levels as 140-350 g per week for females and 210-420 g per week for males ^1^. Alcohol-associated liver disease (ALD) refers to individuals with SLD and high dosage alcohol consumption as higher than 350 g per week and 420 g per week for men and women, respectively ^1^.

The prognosis of SLD varies from a benign course to progression toward steatohepatitis, which can lead to fibrosis, cirrhosis, cirrhosis decompensation and hepatocellular carcinoma (HCC) ^2^. In addition, there are common extrahepatic SLD complications. Alcohol exposure and metabolic risk factors may further exacerbate disease progression. According to the Global Burden of Disease study, there is no safe level of alcohol consumption ^4^. Alcohol cessation is recommended for individuals with SLD, especially in cases of advanced fibrosis or cirrhosis ^1^.

However, current knowledge regarding the effect of alcohol on SLD remains controversial. For example, systematic reviews and meta-analyses have reported a decreased risk of advanced fibrosis among modest alcohol drinkers with SLD ^5,6^. A more recent meta-analysis compared hepatic and extrahepatic outcomes between MASLD and MetALD, and found that patients with MetALD had higher rates of liver-related events, HCC, and extrahepatic cancers ^7^. However, this study did not directly evaluate the effect of alcohol on MASLD or MetALD and included only studies published after the recent re-classification of SLD definition ^1^. Therefore, historical systematic review and meta-analysis studies mainly focused on specific sub-populations of SLD, or based on data from specific period, lacking conclusive evidence in this respect. This study aims to holistically assess the effect of alcohol consumption on the overall SLD populations through systematic review and meta-analysis.

## Materials and Methods

### Search strategy

This systematic review and meta-analysis was conducted in accordance with the PRISMA guidelines ^8^. The study protocol was registered in PROSPERO (CRD420251079136).

We systematically searched EMBASE, MEDLINE, Web of Science, and the Cochrane Central Register of Controlled Trials. In addition, the reference lists of relevant reviews and eligible articles were manually screened to identify additional studies. Key phrases, namely “NAFLD”, “MASLD”, “MetALD”, “steatotic liver disease”, “fatty liver disease” and “alcohol” were used. The full search strategy is described in supplementary file.

Two authors (M.Y. and X.L.) independently screened and selected studies using the Covidence platform. Any discrepancies were discussed and resolved through consensus. A third author adjudicated unresolved disagreements.

### Eligibility criteria

Studies were eligible if they investigated the association between alcohol consumption and progression of SLD in adult patients (≥18 years) and compared outcomes between alcohol consumers and abstainers. SLD had to be diagnosed using imaging modalities, liver biopsy, or validated noninvasive diagnostic methods.

We included all original observational studies, namely cohort, case–control and cross-sectional designs. Studies with pediatric patients (<18 years), insufficient or incomplete data and animal studies were excluded. Studies in which SLD was attributable to secondary causes other than metabolic dysfunction or alcohol consumption were also excluded.

### Alcohol dosage stratification

Alcohol intake was stratified according to the definitions of MASLD, MetALD, and ALD. Mild alcohol consumption was defined based on the MASLD criteria as <30 g/day for men and <20 g/day for women, or <210 g/week for men and <140 g/week for women. Moderate alcohol consumption was defined according to the MetALD criteria as 30–60 g/day for men and 20–50 g/day for women, or 210–420 g/week for men and 140–350 g/week for women. High alcohol consumption was defined in accordance with the ALD criteria as >60 g/day for men and >50 g/day for women, or >420 g/week for men and >350 g/week for women.

Studies that did not report alcohol dosage stratification were also included in the analysis comparing alcohol consumers with abstainers. Alcohol drinkers were defined as patients who consumed any amount of alcohol, including those without a specified dosage level.

### Outcome measures

The primary outcomes included liver related events, defined as fibrosis, cirrhosis, cirrhosis decompensation (ascites, spontaneous bacterial peritonitis, variceal bleeding, hepatic encephalopathy), liver transplantation, HCC, liver related mortality. Secondary outcomes analyzed mortality (all cause and liver-related) and cardiovascular outcomes, including acute myocardial infarction, heart failure, stroke, cardiovascular mortality and arterial hypertension.

Hazard ratio (HR) or odds ratio (OR) with 95% CI between alcohol consumers and nondrinkers were extracted as the effect estimates. Meta-analyses were conducted when at least two studies reported data for a given outcome. A fixed-effect model was used to pool estimates within one study with alcohol dosage categories that differed from our predefined stratification. In addition, a random-effects model was used to pool effect estimates across studies.

### Quality assessment and data extraction

The methodological quality of eligible cohort and case–control studies was assessed using the Newcastle–Ottawa Scale (NOS), while a modified NOS was applied to cross-sectional studies. Quality assessment was performed independently by two reviewers, with disagreements resolved through consensus.

Data extraction was conducted independently by two reviewers (M.Y. and X.L.). Characteristics of the extracted studies included first author, year of publication, country and study design. Study population data included country, study design, sample size, sex distribution, method of alcohol intake assessment, alcohol dosage measurement and method of hepatic steatosis assessment.

### Statistical analysis

Statistical analyses were performed using STATA version 19.5. The heterogeneity level within studies was calculated using *I*^2^ statistics. Consistent with established Cochran Q test, *I*^2^ values <25% were considered to indicate low heterogeneity, 25–50% moderate heterogeneity, and >75% high heterogeneity. A two-sided *p* value <0.05 was considered statistically significant for all analyses. Funnel plot was generated to assess publication asymmetry for potential bias when an outcome included more than 10 studies.

## Results

The literature search identified 2,228 studies. After title and abstract screening, 152 studies were eligible for full-text review. An additional 6 studies were identified through manual searching. Ultimately, 26 studies were included in the meta-analysis ^9-34^.

Overall, 15 cohort studies and 11 cross-sectional studies were included, comprising a total of 466,611 adult patients from 13 countries (Supplementary table 2). The meta-analysis included studies published both before and after the Delphi consensus on SLD definition ^1^, with 7 studies using the MASLD terminology and 19 studies using the NAFLD terminology. Most studies defined SLD based on liver biopsy or imaging modalities, including abdominal ultrasound, transient elastography, and computed tomography. Noninvasive indices were also used, such as the lipid accumulation product (LAP), fatty liver index (FLI), and hepatic steatosis index (HSI). Alcohol intake was assessed using standardized questionnaires, including the AUDIT-C, lifetime drinking history (LDH), or self-reported consumption, as well as laboratory measures such as phosphatidylethanol (PEth) levels or carbohydrate-deficient transferrin (CDT).

Overall, 13 cross-sectional studies and 6 cohort studies evaluated the effect of alcohol consumption on liver-related outcomes in patients with SLD (Figure 2). Among cross-sectional studies, the risk of liver-related outcomes did not differ between all alcohol consumers and abstainers (OR 1.13, 95% CI 0.77–1.65; *p*<0.001, *I*^2^= 86.6%; Figure 2A), nor between different alcohol consumption categories and abstainers. In contrast, three cohort studies reported a higher risk of liver-related events among high-dose alcohol consumers compared with abstainers (HR 2.97, 95% CI 1.61–5.50; *p*<0.001, *I*^2^=91.8%; Figure 2B), as well as among all alcohol consumers compared with abstainers (HR 1.93, 95% CI 1.60–2.33; *p*<0.001, *I*^2^= 93.7%; Figure 2B). Eight cohort studies reported no association between alcohol consumption and overall mortality in analyses without dosage stratification (HR 1.05, 95% CI 0.98–1.14; *p*<0.001, *I*^2^=89.3%; Figure 3). Mortality risk did not differ between moderate or high alcohol consumers and abstainers; however, low alcohol consumption was associated with a reduced risk of mortality (HR 0.88, 95% CI 0.78–0.98; *p*=0.001; *I*^2^=75.5%; Figure 3). Only one study reported liver-related mortality, which was insufficient for meta-analysis.

**Figure 1.**
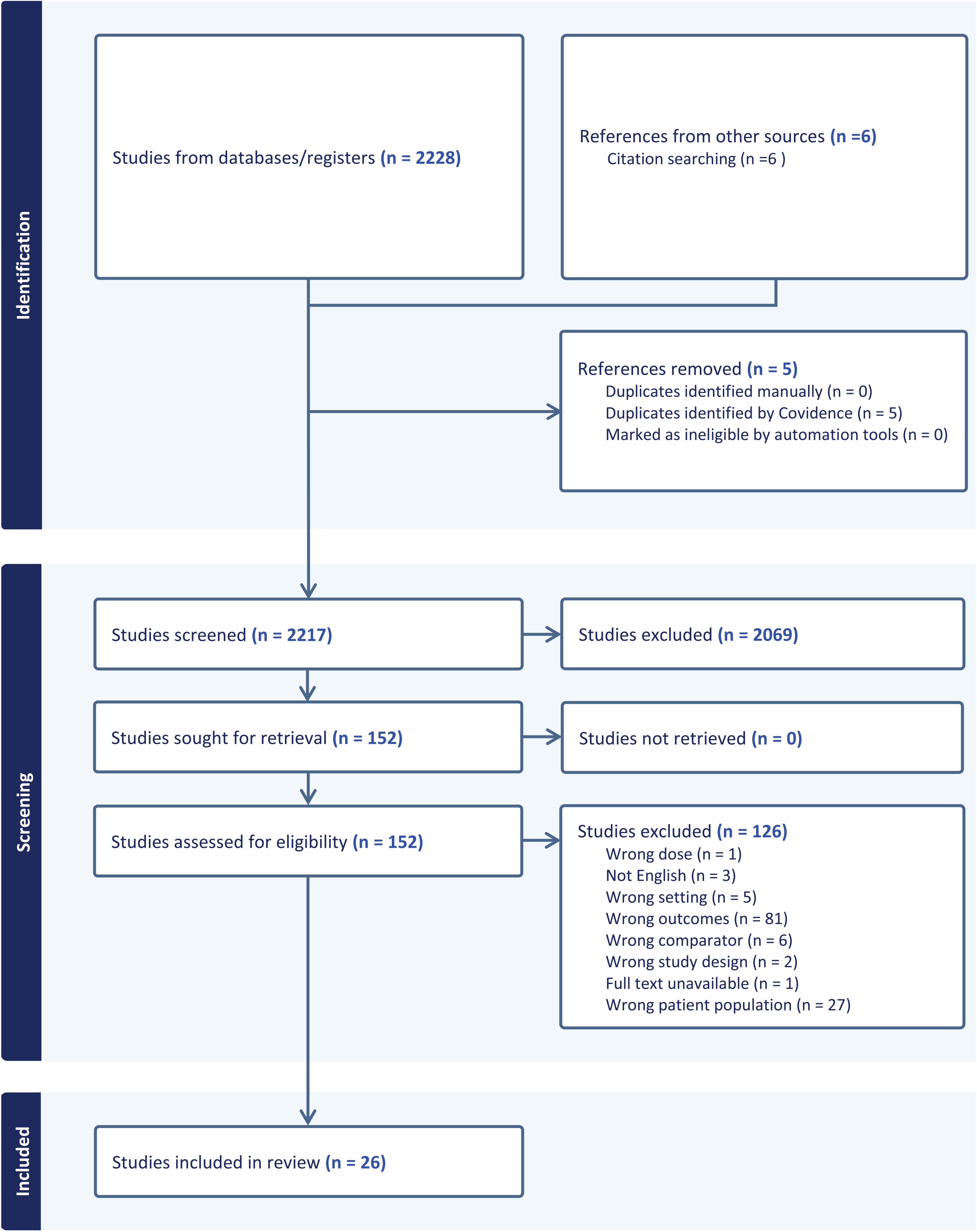
PRISMA flowchart of the study selection.

**Figure 2.**
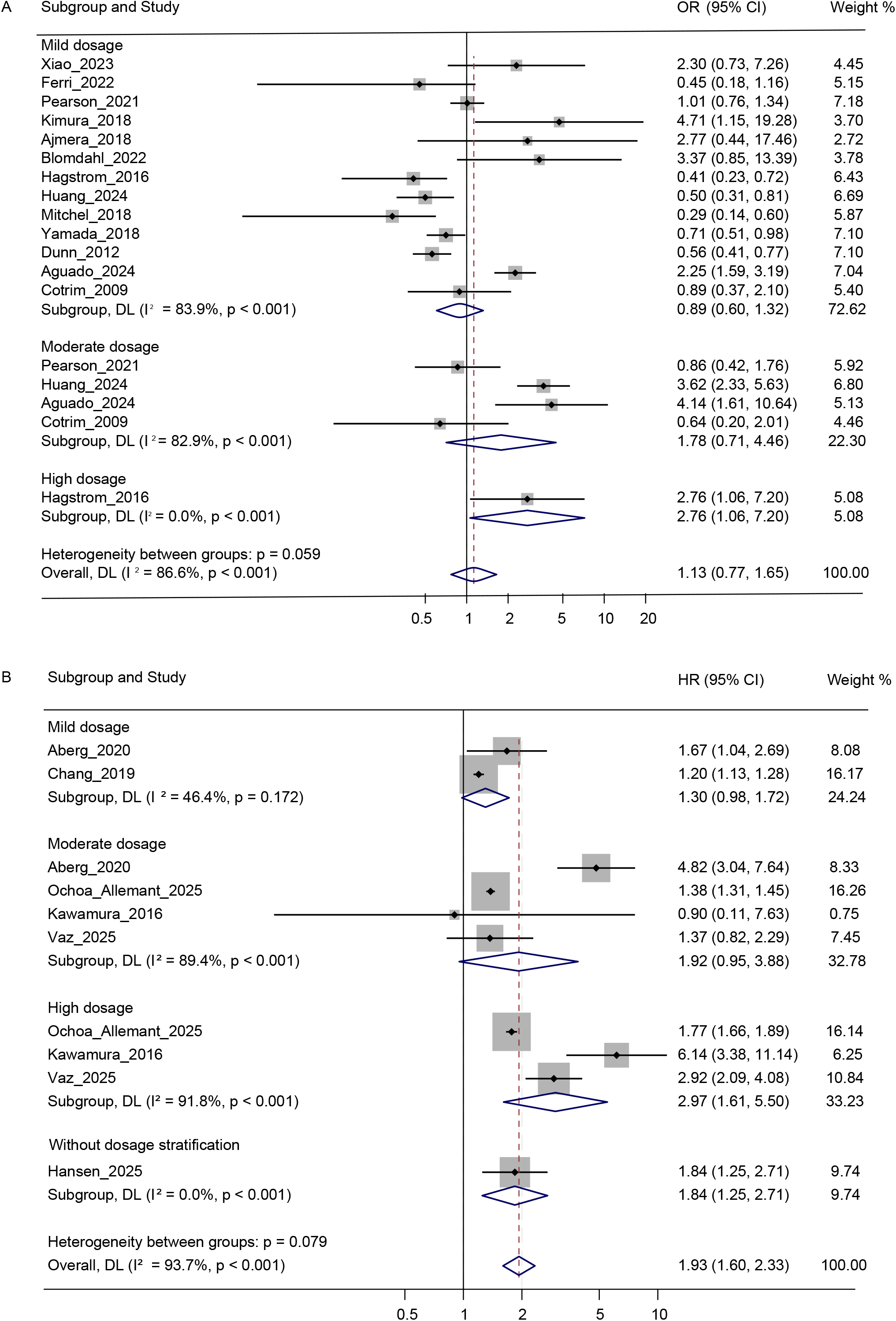
Forest plot of liver-related outcomes stratified by low, moderate, high alcohol consumption and all alcohol consumers, among patients with steatotic liver disease (SLD), in (A) cross sectional studies and (B) cohort studies.

**Figure 3.**
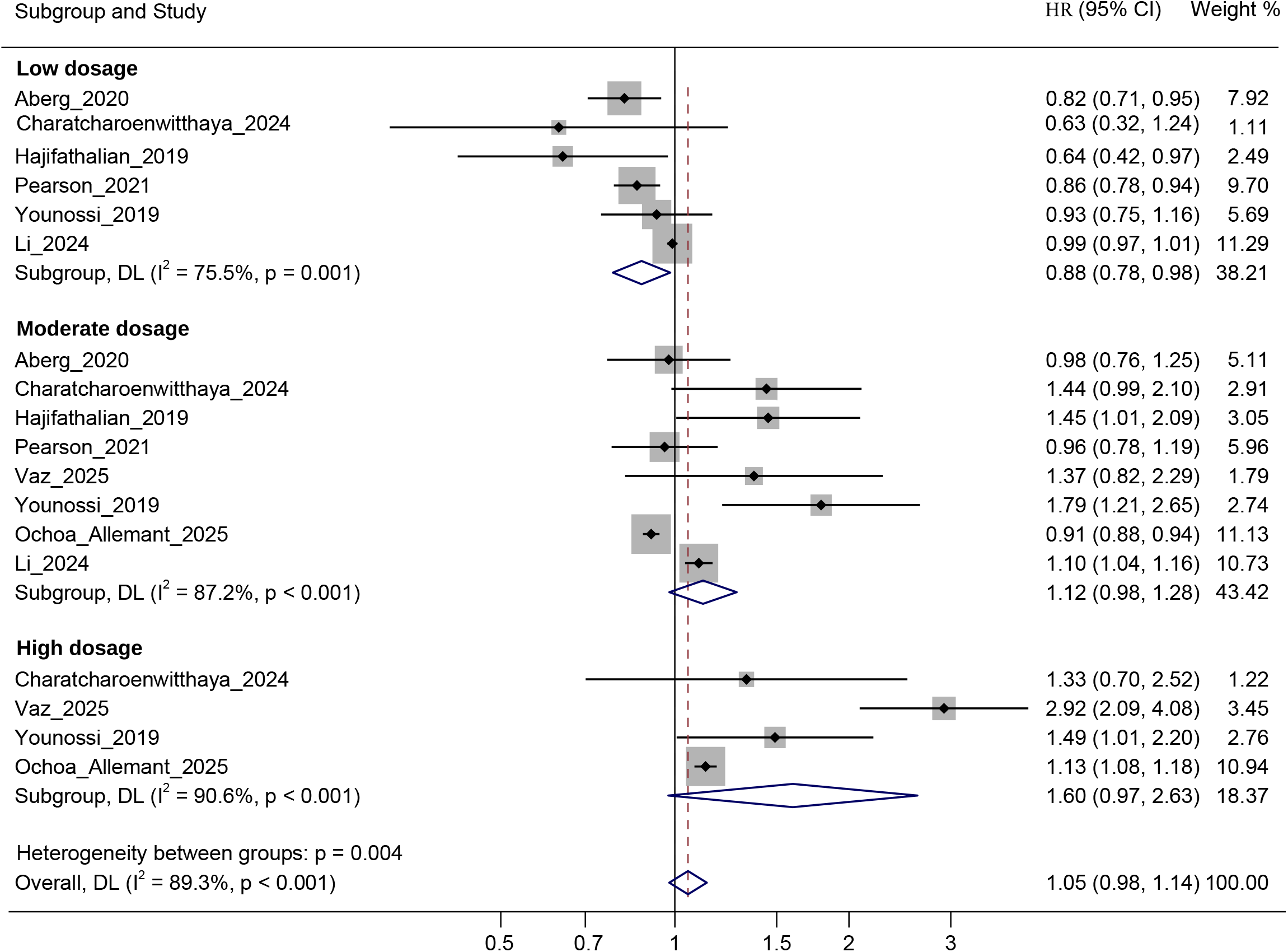
Forest plot of mortality outcomes stratified by low, moderate, high alcohol consumption and all alcohol consumers among patients with SLD.

Three cross-sectional studies examined HCC incidence among alcohol consumers and abstainers with SLD and found no significant difference (OR 1.11, 95% CI 0.44–2.79; *p*=0.025; *I*^2^=72.9%; Figure 4A). Similarly, two cross-sectional studies showed no difference in overall cancer incidence between abstainers and mild alcohol consumers (OR 1.00, 95% CI 0.51–1.95; *p*=0.055; *I*^2^=60.5%; Figure 4B). In contrast, moderate alcohol consumption was associated with a higher cancer incidence compared with abstinence (HR 1.41, 95% CI 1.13–1.78; *p*=0.677; *I*^2^=0.0%; Figure 4C). No significant difference in the risk of cardiovascular diseases as observed between mild alcohol consumers (OR 0.96, 95% CI 0.53–1.74; *p*=0.029; *I*^2^=71.7%; Supplementary figure 1A) or alcohol consumers in general (OR 0.99, 95% CI 0.66–1.49; *p*=0.043; *I*^2^=63.2%; Supplementary Figure S1A) and abstainers in cross-sectional studies. Additionally, two cohort studies reported no difference in cardiovascular risk between moderate alcohol consumers or all alcohol consumers and abstainers (HR 0.83, 95% CI 0.68–1.02; *p*=0.002; *I*^2^=89.9% Figure S1C).

**Figure 4.**
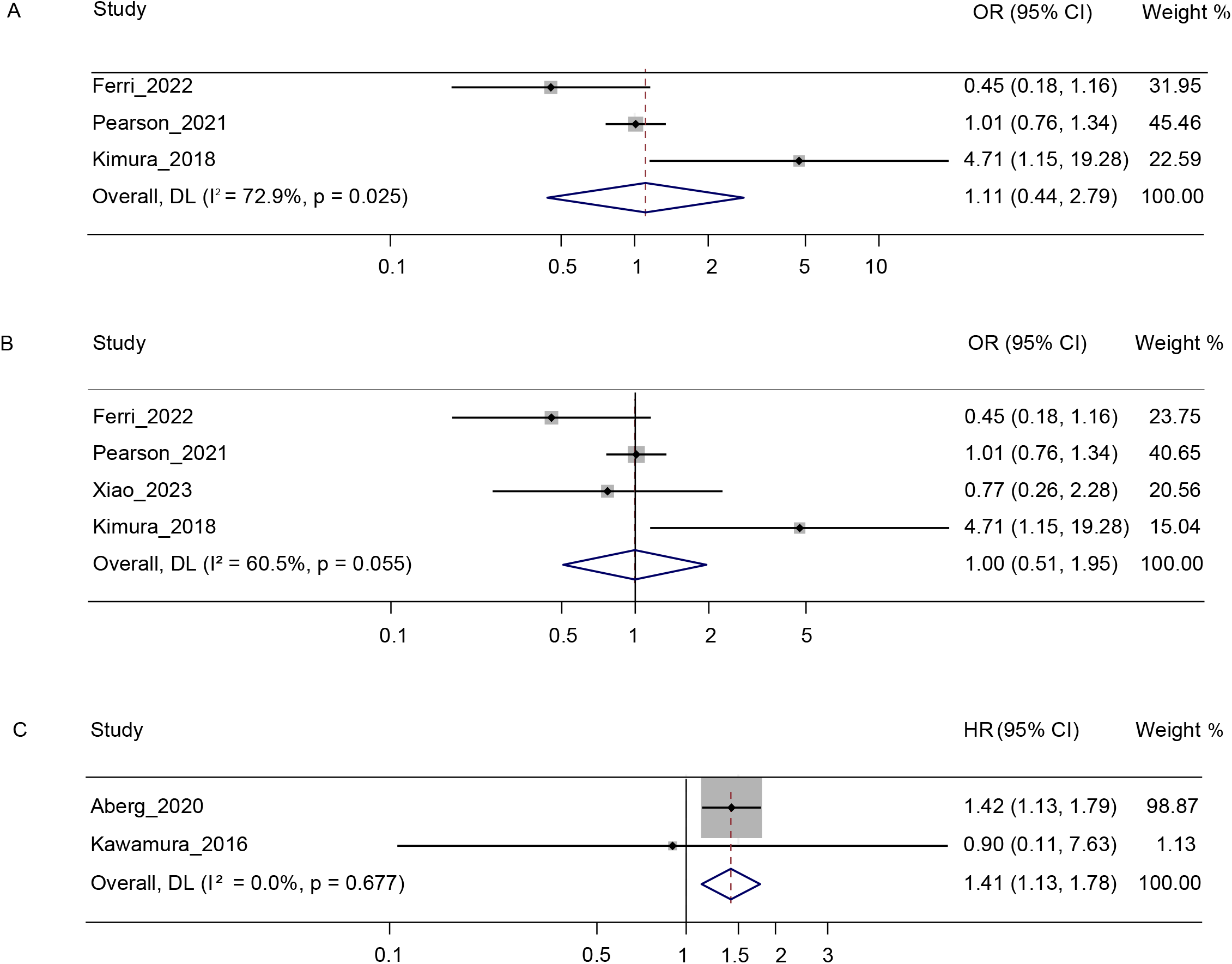
Forest plots of (A) hepatocellular carcinoma (HCC) incidence among patients with steatotic liver disease (SLD) who consume mild amounts of alcohol in cross-sectional studies; (B) overall cancer incidence among patients with SLD who consume mild amounts of alcohol in cross-sectional studies; and (C) overall cancer incidence among patients with SLD who consume moderate amounts of alcohol in cohort studies.

The methodological quality of the included studies was assessed using the Newcastle–Ottawa Scale (NOS) (Supplementary Table 3, 4). In total, 17 cohort studies and nine cross-sectional studies were evaluated. Among cohort studies, 16 were rated as high quality (NOS score 7–9), and one study was of moderate quality (NOS score 4–6). Among cross-sectional studies, four were rated as very good quality, four as good quality, and one as satisfactory quality. No studies were classified as low quality.

## Discussion

In this systematic review and meta-analysis, we comprehensively evaluated the associations of different levels of alcohol consumption with hepatic and extrahepatic outcomes in individuals with SLD. Different from previous studies ^5,6^, this meta-analysis holistically assessed the dose-dependent effects of alcohol on both hepatic and extrahepatic outcomes in SLD, by including studies published both before and after the new definition of SLD terminology. Because definitions of a standard drink vary internationally, alcohol intake categories were harmonized according to MASLD, MetALD, and ALD criteria ^1,35^. Alcohol exposure assessment remains challenging and may be underestimated due to provider-related interviewing limitations or patient-related factors such as social stigma, cognitive impairment, or language barriers ^35^. Because some studies did not report alcohol intake by dosage, all available studies were combined in an additional analysis to evaluate the general effect of alcohol consumption on SLD, independent of dose categorization.

The outcomes included liver-related events, malignancy, cardiovascular disease, and all-cause mortality. We found that high-dose alcohol consumption, as well as overall alcohol intake, was associated with an increased risk of liver-related events in cohort studies. Mechanistically, it is known that high-dose alcohol intake promotes oxidative stress and mitochondrial dysfunction, resulting in hepatic inflammation and fibrosis progression ^36^. Only one cross-sectional study reported high-dose alcohol consumption, which was insufficient for quantitative synthesis ^9^. Subgroup analyses of steatohepatitis and fibrosis progression did not show differences between drinkers and abstainers, which does not support previously suggested hepatoprotective effects of low alcohol consumption in NAFLD/MASLD ^10,11,35^.

Moderate alcohol consumption was associated with a higher incidence of malignancy in cohort studies. Alcohol may promote carcinogenesis through DNA adduct formation, which can lead to point mutations ^37^. Although alcohol intake is associated with several cancer types including HCC, breast, colorectal, laryngeal, esophageal, and oral cancers, most included studies in the analysis did not specify cancer types ^38^. Carcinogenic effects were observed only with moderate alcohol consumption in cohort studies; however, the number of cohort studies evaluating mild and moderate alcohol intake was insufficient to allow reliable stratified analysis. Furthermore, no association was found between mild alcohol consumption and HCC incidence in the meta-analysis, despite previous reports suggesting a hepatocarcinogenic effect of mild alcohol intake in patients with NAFLD ^12^. Therefore, additional studies are needed to clarify the carcinogenic potential of different levels of alcohol consumption in individuals with SLD.

No significant differences in cardiovascular outcomes were observed between mild alcohol drinkers or overall drinkers and abstainers with SLD in cross-sectional analyses. These findings do not support previously reported cardiovascular benefits of modest alcohol consumption, attributed to lower rates of carotid plaque formation, ischemic events, and higher HDL levels ^13,14^. According to the meta-analysis, mild alcohol consumption was associated with lower all-cause mortality compared with abstinence in patients with SLD in cohort studies, whereas no significant differences were observed between abstainers and moderate or high alcohol consumers. Among the eight cohort studies reporting mortality outcomes, only one study reported liver-related mortality and one reported cardiovascular mortality, while the remaining studies did not specify cause of death. In NAFLD cohorts, the most common causes of death among abstainers and mild drinkers are cardiovascular disease, followed by extrahepatic malignancy and liver-related mortality ^39^. The lower rate of all-cause mortality observed in mild drinkers has often been attributed to potential cardioprotective effects of low-dose alcohol consumption ^13,14^, but it is highly questionable whether mild alcohol consumption confers a true protective effect. Further research is needed to carefully investigate this question, ideally by examining cause-specific mortality and accounting for comorbidities, socioeconomic status and lifestyle factors ^40^. Given that extrahepatic malignancies are the second most common cause of death in individuals with SLD, future studies should also consider cancer detection timing, treatment response, and cancer subtype ^35^.

This study has several limitations. First, heterogeneity may have been introduced by differences in the assessment of steatosis, liver-related outcomes, and alcohol intake. Second, the hepatic and extrahepatic effects of alcohol may vary by fibrosis stage, degree of steatosis, age, sex, genetic background, socioeconomic factors, and comorbidities. Limited available data precluded subgroup analyses based on these modifiers ^41^, potentially contributing to the substantial statistical heterogeneity observed across studies. Third, the observed association between low-dose alcohol consumption and reduced all-cause mortality remains unexplained and may reflect residual confounding; further studies are needed to determine whether this represents a true causal relationship. Finally, because all included studies were observational, causal inferences cannot be made. Reverse causation is also possible, as low-dose daily alcohol use may itself promote hepatic steatosis ^35^.

Despite these limitations, this study has notable strengths. We included studies published both before and after the Delphi consensus to comprehensively capture SLD phenotypes. Alcohol exposure was standardized using MASLD, MetALD, and ALD thresholds, reducing variability in dose classification. Cohort and cross-sectional studies were analyzed separately to account for differences in temporality, effect measures, and risk of bias. Only studies with confirmed hepatic steatosis were included, and ALD studies without documented steatosis were excluded, given that steatosis is present in ∼90% of ALD cases ^42^. Finally, we examined both hepatic and extrahepatic outcomes to provide a broader clinical perspective.

In conclusion, this meta-analysis indicates that high-dose alcohol intake increases the risk of liver-related complications in individuals with SLD, while moderate intake may elevate overall cancer risk. Further studies are needed to clarify dose–response relationships and the influence of metabolic, genetic, and environmental modifiers on alcohol-related outcomes in SLD.

## Data Availability

All data are available from the main manuscript and supplementary file.

## A list of abbreviations in the order of appearance

SLD: steatotic liver disease
MASLD: metabolic dysfunction associated with steatotic liver disease
NAFLD: non-alcoholic fatty liver disease
MetALD: metabolic dysfunction associated alcohol related liver disease
ALD: alcohol associated liver disease
HCC: hepatocellular carcinoma
HR: Hazard ratio
OR: odds ratio
NOS: Newcastle–Ottawa Scale
LAP: lipid accumulation product
FLI: fatty liver index
HSI: hepatic steatosis index
AUDIT-C: alcohol use disorders identification test
LDH: lifetime drinking history
PEth: phosphatidylethanol
CDT: carbohydrate-deficient transferrin
HDL: high-density lipoprotein

## Acknowledgment

The authors would like to thank Dr. Maarten Engel from the Erasmus MC Medical Library for developing the search strategies.

