## Supplementary information for "Holistic assessment of the effect of alcohol consumption on steatotic liver disease: systematic review and meta-analysis"

| **Section and Topic** | **Item #** | **Checklist item** | **Location where item is reported** |
| --- | --- | --- | --- |
| **TITLE** | | |  |
| Title | 1 | Identify the report as a systematic review. | 1 |
| **ABSTRACT** | | |  |
| Abstract | 2 | See the PRISMA 2020 for Abstracts checklist. | 2 |
| **INTRODUCTION** | | |  |
| Rationale | 3 | Describe the rationale for the review in the context of existing knowledge. | 3 |
| Objectives | 4 | Provide an explicit statement of the objective(s) or question(s) the review addresses. | 3 |
| **METHODS** | | |  |
| Eligibility criteria | 5 | Specify the inclusion and exclusion criteria for the review and how studies were grouped for the syntheses. | 4 |
| Information sources | 6 | Specify all databases, registers, websites, organisations, reference lists and other sources searched or consulted to identify studies. Specify the date when each source was last searched or consulted. | 4 |
| Search strategy | 7 | Present the full search strategies for all databases, registers and websites, including any filters and limits used. | Supplementary file |
| Selection process | 8 | Specify the methods used to decide whether a study met the inclusion criteria of the review, including how many reviewers screened each record and each report retrieved, whether they worked independently, and if applicable, details of automation tools used in the process. | 4 |
| Data collection process | 9 | Specify the methods used to collect data from reports, including how many reviewers collected data from each report, whether they worked independently, any processes for obtaining or confirming data from study investigators, and if applicable, details of automation tools used in the process. | 4 |
| Data items | 10a | List and define all outcomes for which data were sought. Specify whether all results that were compatible with each outcome domain in each study were sought (e.g. for all measures, time points, analyses), and if not, the methods used to decide which results to collect. | 4 |
|  | 10b | List and define all other variables for which data were sought (e.g. participant and intervention characteristics, funding sources). Describe any assumptions made about any missing or unclear information. | 4 |
| Study risk of bias assessment | 11 | Specify the methods used to assess risk of bias in the included studies, including details of the tool(s) used, how many reviewers assessed each study and whether they worked independently, and if applicable, details of automation tools used in the process. | 5 |
| Effect measures | 12 | Specify for each outcome the effect measure(s) (e.g. risk ratio, mean difference) used in the synthesis or presentation of results. | 4 |
| Synthesis methods | 13a | Describe the processes used to decide which studies were eligible for each synthesis (e.g. tabulating the study intervention characteristics and comparing against the planned groups for each synthesis (item #5)). | 4 |
|  | 13b | Describe any methods required to prepare the data for presentation or synthesis, such as handling of missing summary statistics, or data conversions. | 4 |
|  | 13c | Describe any methods used to tabulate or visually display results of individual studies and syntheses. | 4 |
|  | 13d | Describe any methods used to synthesize results and provide a rationale for the choice(s). If meta-analysis was performed, describe the model(s), method(s) to identify the presence and extent of statistical heterogeneity, and software package(s) used. | 5 |
|  | 13e | Describe any methods used to explore possible causes of heterogeneity among study results (e.g. subgroup analysis, meta-regression). | 5 |
|  | 13f | Describe any sensitivity analyses conducted to assess robustness of the synthesized results. | 4 |
| Reporting bias assessment | 14 | Describe any methods used to assess risk of bias due to missing results in a synthesis (arising from reporting biases). | 5 |
| Certainty assessment | 15 | Describe any methods used to assess certainty (or confidence) in the body of evidence for an outcome. | N/A |
| **RESULTS** | | |  |
| Study selection | 16a | Describe the results of the search and selection process, from the number of records identified in the search to the number of studies included in the review, ideally using a flow diagram. | 5 |
|  | 16b | Cite studies that might appear to meet the inclusion criteria, but which were excluded, and explain why they were excluded. | Figure 1 |
| Study characteristics | 17 | Cite each included study and present its characteristics. | Supplementary table 2 |
| Risk of bias in studies | 18 | Present assessments of risk of bias for each included study. | Supplementary table 3 A, B |
| Results of individual studies | 19 | For all outcomes, present, for each study: (a) summary statistics for each group (where appropriate) and (b) an effect estimate and its precision (e.g. confidence/credible interval), ideally using structured tables or plots. | Figure 2-4, Supplementary figure 1-3 |
| Results of syntheses | 20a | For each synthesis, briefly summarise the characteristics and risk of bias among contributing studies. | Supplementary table 2, 3 |
|  | 20b | Present results of all statistical syntheses conducted. If meta-analysis was done, present for each the summary estimate and its precision (e.g. confidence/credible interval) and measures of statistical heterogeneity. If comparing groups, describe the direction of the effect. | Figure 2-4, Supplementary figure 1-3 |
|  | 20c | Present results of all investigations of possible causes of heterogeneity among study results. | 8 |
|  | 20d | Present results of all sensitivity analyses conducted to assess the robustness of the synthesized results. | 6 |
| Reporting biases | 21 | Present assessments of risk of bias due to missing results (arising from reporting biases) for each synthesis assessed. | N/A |
| Certainty of evidence | 22 | Present assessments of certainty (or confidence) in the body of evidence for each outcome assessed. | N/A |
| **DISCUSSION** | | |  |
| Discussion | 23a | Provide a general interpretation of the results in the context of other evidence. | 7 |
|  | 23b | Discuss any limitations of the evidence included in the review. | 8 |
|  | 23c | Discuss any limitations of the review processes used. | 8 |
|  | 23d | Discuss implications of the results for practice, policy, and future research. | 9 |
| **OTHER INFORMATION** | | |  |
| Registration and protocol | 24a | Provide registration information for the review, including register name and registration number, or state that the review was not registered. | 3 |
|  | 24b | Indicate where the review protocol can be accessed, or state that a protocol was not prepared. | PROSPERO |
|  | 24c | Describe and explain any amendments to information provided at registration or in the protocol. | No amendments |
| Support | 25 | Describe sources of financial or non-financial support for the review, and the role of the funders or sponsors in the review. | N/A |
| Competing interests | 26 | Declare any competing interests of review authors. | 2 |
| Availability of data, code and other materials | 27 | Report which of the following are publicly available and where they can be found: template data collection forms; data extracted from included studies; data used for all analyses; analytic code; any other materials used in the review. | N/A |

*From:* Page MJ, McKenzie JE, Bossuyt PM, Boutron I, Hoffmann TC, Mulrow CD, et al. The PRISMA 2020 statement: an updated guideline for reporting systematic reviews. BMJ 2021;372:n71. doi: 10.1136/bmj.n71. This work is licensed under CC BY 4.0. To view a copy of this license, visit <https://creativecommons.org/licenses/by/4.0/>

Supplementary Table 1. PRISMA 2020 checklist

| # | Study | Country | Study design | Patient number | Method of assessing alcohol | Alcohol dosage measurement | Hepatic steatosis measurement |
| --- | --- | --- | --- | --- | --- | --- | --- |
| 1 | Aberg, 2020 | Finland | Retrospective cohort | 8345 | Interview | 0-9, 10-19, 20-29, 30-39, and 40-49 g/day | FLI≥ 60 |
| 2 | Ajmera 2018 | USA | Cohort | 285 | AUDIT C questionnaire | ≤2 drinks/day- modest drinker | Biopsy |
| 3 | Blomdahl 2022 | Sweden | Retrospective cohort | 82 | AUDIT C/ interview/ PEth | 0–2.99 g/week; 3–65.99 g/week; ≥ 66 g/week | Imaging |
| 4 | Chang 2019 | Korea | Cohort | 58927 | Questionnaire | Light: 1- 10g/day women, 1- 10g/day- man  Moderate drinking- 10-20 g/day- women, 10-30- men | Abdominal ultrasound |
| 5 | Charatcharoenwitthaya, 2024 | Thailand | Retrospective cohort | 8304 | Self reported questionnaire | Abstinence/light < 10g/day, moderate 10- 20 (women), < 30 (men),  Risk drinking 20-50g/day women, 30-60g/day men,  Heavy drinking >50g/day women, >60g/day men | LAP score≥ 30,5 men, ≥23,0 women |
| 6 | Ferri 2022 | Italy | Cross sectional | 276 | Interview | Abstainers<1g/week,  Low consumers 1-70g/week  Moderate consumers 71-210 g/week men and 71-140 g/ week women | Abdominal ultrasound |
| 7 | Hagstrom 2016 | Sweden | Cross sectional | 120 | LDH, AUDIT, PEth | 0-3.6 g/week, 3.6-12 g/week, 12-37.2 g/week, 37.2-159.6 g/week  PEth value < 0.3 micromol/L, >0,3 micromol/L | Liver biopsy |
| 8 | Hajifathalian 2019 | USA | Prospective cohort | 4568 | Interview | <7 g/day- nondrinkers  7-21g/day- modest alcohol consumption  ≥21g/day- high | HSI >36 |
| 9 | Hara 2019 | Japan | Cross sectional | 1190 | Interview | Light drinking: 0- 20g | Abdominal ultrasound |
| 10 | Huang 2024 | China | Retrospective cross sectional | 713 | AUDIT | Mild-moderate: 1-20g/day, Excessive>20g/day women  Nondrinking-0g/day, mild-moderate: 1-<30g/day, Excessive>30g/day men | Abdominal ultrasound/ CT/ MRI |
| 11 | Mitchel 2018 | Australia | Cross sectional observational | 187 | Interview | Nondrinking-0, <70g/week, >70g/week | Liver biopsy |
| 12 | Pearson 2021 | USA | Retrospective cohort | 6622 | AUDIT C questionnaire | AUDIT C: 1-3 in men, 1-2 in women low level drinking,  4-12 men, 3-12 women -unhealthy drinking | Cirrhosis, diagnosis of exclusion |
| 13 | Sinn 2014 | South Korea | Cross sectional | 2280 | Questionnaire | Mild <20 g/day | Abdominal ultrasound |
| 14 | Vaz 2025 | Sweden | Prospective cohort | 7499 | PEth | No stratification | Collected from ICD medical history |
| 15 | Xiao 2023 | Singapore | Cross sectional | 4723 | Interview | Modest drinker < 21/week standard drink for men, 14 standard / week for women | Transient elastography CAP> 248dB/m |
| 16 | Yamada 2018 | Japan | Cross sectional | 178 | Self reported questionnare | Nondrinker, <20g/day - light alcohol consumption | Liver biopsy |
| 17 | Younossi 2019 | USA | Cross sectional | 4264 | Questionnaire | Men: minimum drinker: 0-30 g/week, moderate> 30g/week, but < 20/day, substantially: 20-30g/day, Excessive> 30g/day  women: minimum drinker: 0-15 g/week, moderate> 15g/week, but < 10/day, substantially: 10-15 g/day, Excessive> 15g/day  Binge drinking> 50 g/day | Abdominal ultrasound |
| 18 | Dunn 2012 | USA | Cross sectional | 582 | AUDIT | <20g/day - modest alcohol consumption | Liver biopsy |
| 19 | Aguado 2024 | Spain | Prospective cohort | 2303 | Questionnaire | 0-40 g/ week- very low, 50-90 g/ week- low, Moderate: women: 100-130g/week,  100-200 g/week men, MetALD- women: 140-350g/week, 210-420 g/ week men | Transient elastography CAP> 275 dB/m |
| 20 | Kimura 2018 | Japan | Prospective cohort | 301 | Self reported questionnaire | Mild drinker- <20g/day | Abdominal ultrasound |
| 21 | Ochoa-Allemant 2025 | USA | Prospective cohort | 341601 | Interview | <20g/day, 20-40g/day | Biopsy |
| 22 | Kawamura 2016 | Japan | Retrospective cohort | 9959 | Questionnaire | NAFLD<20/day  Low intermediate 20-39 g/day  High intermediate 40-69 g/day  >70g - AFLD | Abdominal ultrasound |
| 23 | Cotrim 2009 | Brazil | Cross sectional | 132 | Interview | Nondrinker, <20g/day, 20-40g/day | Biopsy |
| 24 | Hansen 2025 | Denmark | Retrospective cohort | 192 | Interview/ PEth/CDT | 100 g/day | Histology |
| 25 | Li 2024 | China | Retrospective cohort | 2408 | No information | MASLD, MetALD criteria | Abdominal ultrasound |
| 26 | VanWagner 2017 | USA | Prospective cohort | 570 | Interview | No stratification | CT |

Supplementary Table 2. Characteristics of the studies included in the meta analysis. PEth=Phosphatidylethanol. LDH=Lifetime drinking history. CDT= Carbohydrate deficient transferrin. NAFLD= Non-alcoholic fatty liver disease. AFLD= Alcoholic fatty liver disease. MASLD=Metabolic-dysfunction associated

steatotic liver disease. MetALD= Metabolic Dysfunction-Associated and Alcohol-Associated Liver Disease. CT= Computed tomography. MRI= Magnetic resonance imaging. CAP= Controlled attenuation parameter. FLI= Fatty liver index. HSI =hepatic steatosis index. ICD= International classification of diseases. AUDIT-C= Alcohol use disorder identification test- consumption

| Study, year | Selection (0-4) | Comparability (0-2) | Outcome (0-3) | Total score (0-9) | Quality |
| --- | --- | --- | --- | --- | --- |
| Aberg, 2020 | ★★★★ | ★★ | ★★ | 8 | High |
| Ajmera 2018 | ★★★ | ★★ | ★★ | 7 | High |
| Blomdahl 2022 | ★★★★ | ★★ | ★★ | 8 | High |
| Chang 2019 | ★★★★ | ★★ | ★★★ | 9 | High |
| Charatcharoenwitthaya, 2024 | ★★★★ | ★★ | ★★ | 8 | High |
| Hajifathalian 2019 | ★★★★ | ★★ | ★★ | 8 | High |
| Pearson 2021 | ★★★ | ★★ | ★★ | 7 | High |
| Vaz 2025 | ★★★ | ★★ | ★★★ | 8 | High |
| Younossi 2019 | ★★★ | ★★ | ★★ | 7 | High |
| Aguado 2019 | ★★★ | ★★ | ★ | 6 | Moderate |
| Kimura 2018 | ★★★ | ★★ | ★★★ | 8 | High |
| Ochoa-Allemant 2025 | ★★★★ | ★★ | ★★★ | 9 | High |
| Kawamura 2016 | ★★★★ | ★★ | ★★★ | 9 | High |
| Hansen 2025 | ★★★★ | ★★ | ★★★ | 9 | High |
| Li 2024 | ★★★★ | ★★ | ★★★ | 9 | High |
| VanWagner | ★★★★ | ★★ | ★★ | 8 | High |
| Younossi 2019 | ★★★★ | ★★ | ★★ | 8 | High |

Supplementary Table 3. Quality assessment by Newcastle Ottawa Scale (NOS) (cohort studies). Quality interpretation: 1-3 points- low, 4-6 points- moderate, 7-9 points- high.

| Study, year | Selection (0-5) | Comparability (0-2) | Outcome (0-3) | Total score (0-10) | Quality |
| --- | --- | --- | --- | --- | --- |
| Xiao 2023 | ★★★★ | ★★ | ★★★ | 9 | Very good |
| Yamada 2018 | ★★★ | ★★ | ★★★ | 8 | Good |
| Ferri 2022 | ★★★★ | ★★ | ★★★ | 9 | Very good |
| Hagstrom 2016 | ★★★ | ★★ | ★★★ | 8 | Good |
| Hara 2019 | ★★★★★ | ★★ | ★★★ | 10 | Very good |
| Mitchell 2018 | ★★★ | ★★ | ★★★ | 8 | Good |
| Sinn 2014 | ★★★★ | ★★ | ★★★ | 9 | Very good |
| Dunn 2012 | ★★★ | ★★ | ★★★ | 8 | Good |
| Cotrim 2009 | ★★ |  | ★★★ | 5 | Satisfactory |

Supplementary Table 4. Quality assessment by Newcastle Ottawa Scale (NOS) (cross sectional studies).

Quality interpretation: 7-8 points- good, 5-6 points –satisfactory, 0-4 points- unsatisfactory.

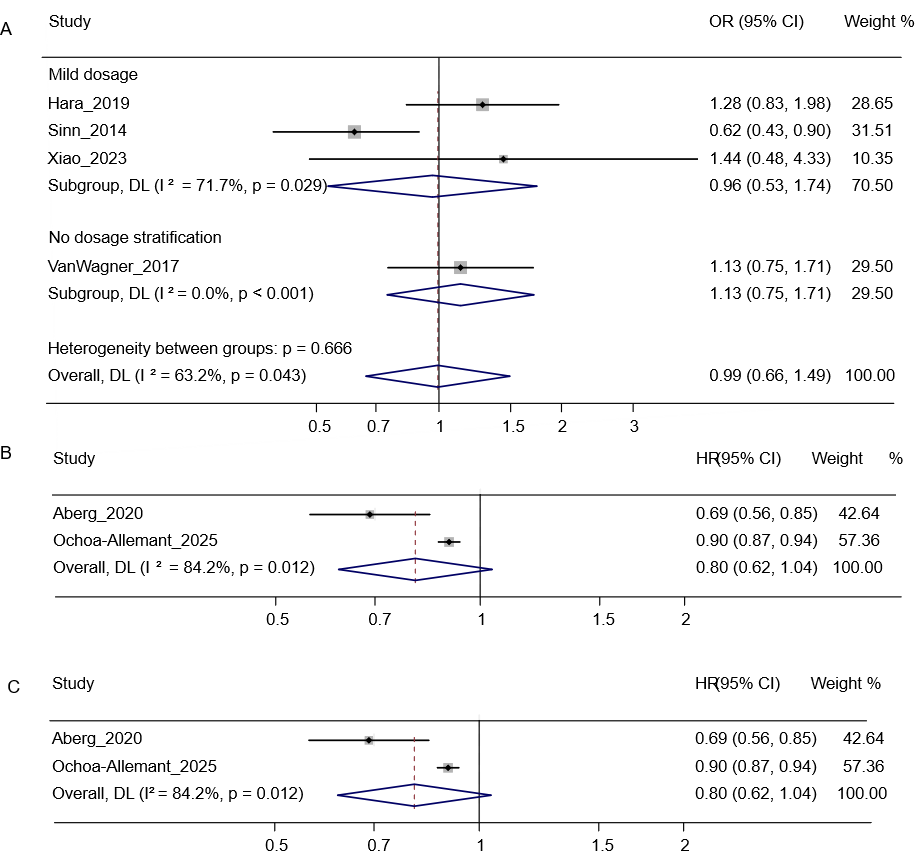

Supplementary figure 1. Forest plot of the risk of cardiovascular disease among patients with steatotic liver disease (SLD): (A) cross-sectional studies; (B) cohort studies stratified by moderate alcohol dosage; and (C) all alcohol consumers in cohort studies

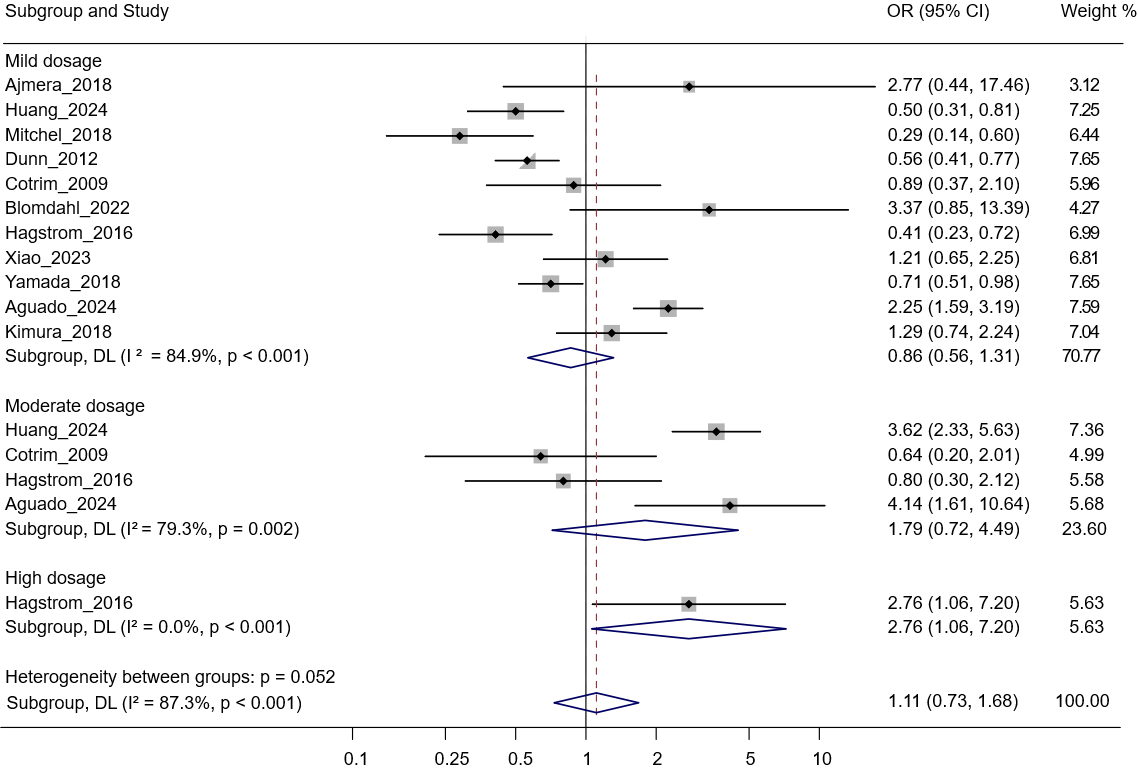

Supplementary figure 2. Forest plot of the fibrosis stratified by low, moderate, high alcohol consumption and all alcohol consumers among patients with steatotic liver disease (SLD) in cross sectional studies

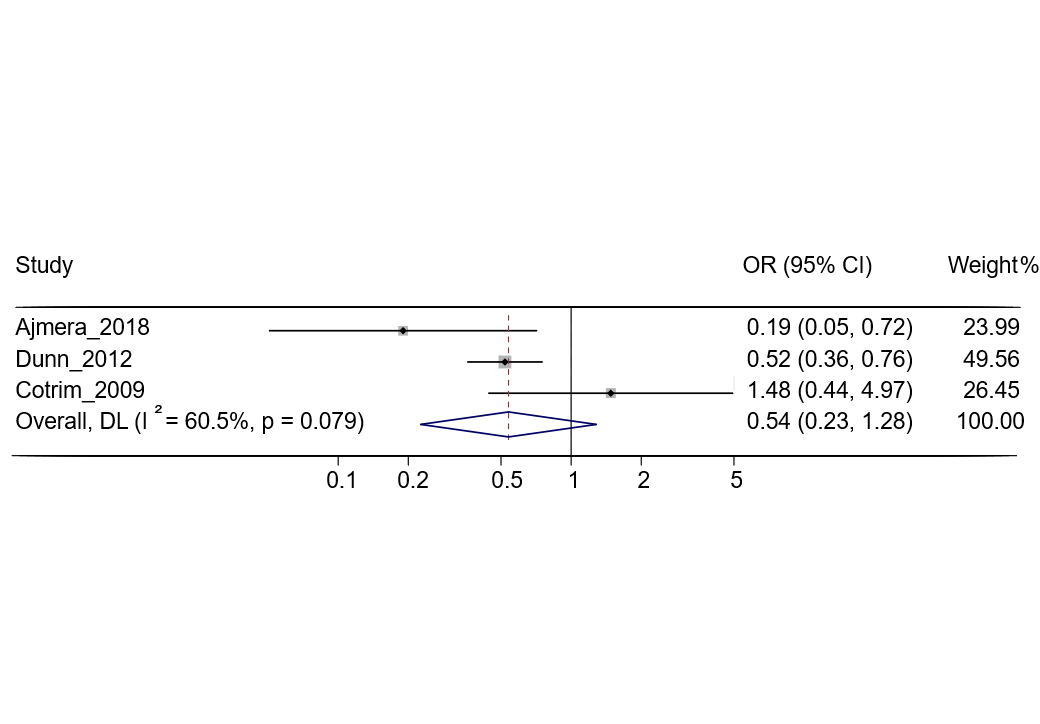

Supplementary figure 3. Forest plot of the steatohepatitis among patients with SLD and mild alcohol drinkers in cross sectional studies

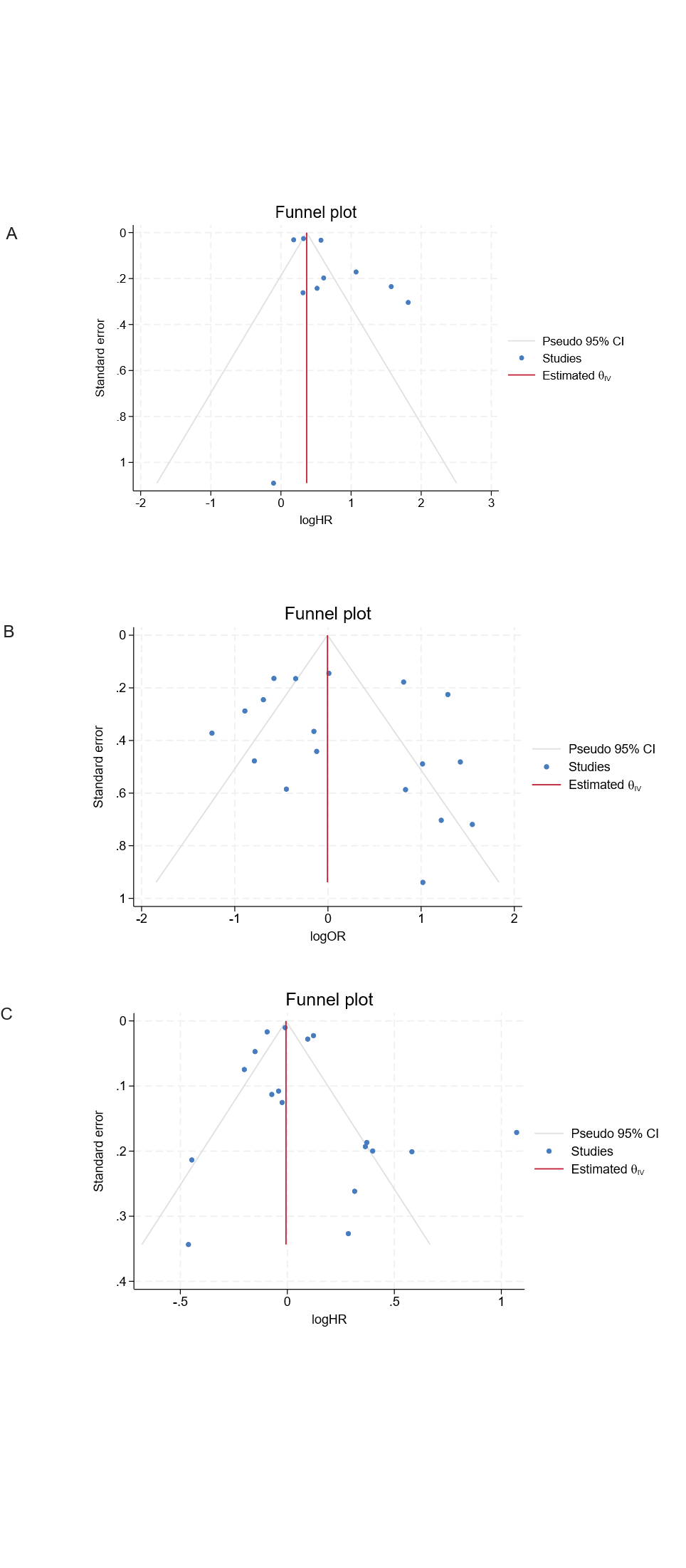

Supplementary figure 4. Funnel plot of liver- related outcomes in (A) cohort studies, (B) cross-sectional studies and (C) mortality

**Full search strategy**

20250603 Madina Yerezhepbayeva

alcohol consumption fatty liver disease

SR

research question: The effects of alcohol consumption on the patients with fatty liver disease, clinical studies only

| **Database searched** | **Platform** | **Years of coverage** | **Records** | **Records after duplicates removed** |
| --- | --- | --- | --- | --- |
| Medline ALL | Ovid | 1946 - Present | 311 | 308 |
| Embase | Embase.com | 1971 - Present | 1414 | 1190 |
| Web of Science Core Collection* | Web of Knowledge | 1975 - Present | 517 | 254 |
| Cochrane Central Register of Controlled Trials | Wiley | 1992 - Present | 430 | 333 |
| Additional Search Engines: Google Scholar** | | | 200 | 137 |
| **Total** | | | **2872** | **2222** |

*Science Citation Index Expanded (1975-present) ; Social Sciences Citation Index (1975-present) ; Arts & Humanities Citation Index (1975-present) ; Conference Proceedings Citation Index- Science (1990-present) ; Conference Proceedings Citation Index- Social Science & Humanities (1990-present) ; Emerging Sources Citation Index (2005-present)

**Google Scholar was searched via "Publish or Perish" to download the results in EndNote.

*Excluded publication types were conference abstracts and case reports*

*The search was limited to the English language and to human studies as documented in the search strategy.*

*The search was limited to exclude children-only studies as documented in the search strategy.*

**Embase 1414**

('nonalcoholic fatty liver'/exp OR 'metabolic fatty liver'/de OR 'metabolic dysfunction associated steatotic liver disease'/de OR 'fatty liver'/de OR Steatohepatitis/de OR (NAFLD OR MAFLD OR MASLD OR METALD OR FLD OR SLD OR MASH OR NASH OR ((nonalcoholic OR non-alcoholic OR metabol*) NEAR/6 (fatty-liver* OR steatotic-liver* OR liver-steatos* OR hepat*-steatos* OR alcoholic-liver* OR liver-fibros* OR hepatic-fibros*)) OR ((fatty-liver* OR steato*) NEAR/3 (diseas*)) OR Steatohepatit* OR ((fatty-liver* OR steatotic-liver* OR liver-steatos* OR hepat*-steatos* OR alcoholic-liver* OR liver-fibros* OR hepatic-fibros*) AND (hepatocellular-carcinom* OR HCC))):ab,ti,kw OR ((metabol*) AND (fatty-liver* OR steatotic-liver* OR hepatic-steatos* OR alcoholic-liver* OR liver-fibros* OR hepatic-fibros* OR Steatohepatit*)):ti) **AND** ('alcohol consumption'/de OR 'alcohol abuse'/exp OR 'drinking behavior'/de OR (((alcohol) NEAR/3 (consumpt* OR intake OR use OR abuse OR drink*)) OR ((drink*) NEAR/3 (behav* OR binge OR heavy))):ab,ti,kw) **AND** ('randomized controlled trial'/de OR 'controlled clinical trial'/de OR 'Controlled clinical trial'/exp OR 'Crossover procedure'/de OR 'Double-blind procedure'/de OR 'Single-blind procedure'/de OR (random* OR factorial* OR crossover* OR (cross NEXT/1 over*) OR placebo* OR ((doubl* OR singl*) NEXT/1 blind*) OR assign* OR allocat* OR volunteer* OR trial):ab,ti,kw OR random*:ti,ab,tt OR 'randomization'/de OR 'intermethod comparison'/de OR placebo:ti,ab,tt OR (compare:ti,tt OR compared:ti,tt OR comparison:ti,tt) OR ((evaluated:ab OR evaluate:ab OR evaluating:ab OR assessed:ab OR assess:ab) AND (compare:ab OR compared:ab OR comparing:ab OR comparison:ab)) OR (open NEXT/1 label):ti,ab,tt OR ((double OR single OR doubly OR singly) NEXT/1 (blind OR blinded OR blindly)):ti,ab,tt OR 'double blind procedure'/de OR (parallel NEXT/1 group*):ti,ab,tt OR (crossover:ti,ab,tt OR 'cross over':ti,ab,tt) OR ((assign* OR match OR matched OR allocation) NEAR/6 (alternate OR group OR groups OR intervention OR interventions OR patient OR patients OR subject OR subjects OR participant OR participants)):ti,ab,tt OR (assigned:ti,ab,tt OR allocated:ti,ab,tt) OR (controlled NEAR/8 (study OR design OR trial)):ti,ab,tt OR (volunteer:ti,ab,tt OR volunteers:ti,ab,tt) OR 'human experiment'/de OR trial:ti,tt ) NOT ((((random* NEXT/1 sampl* NEAR/8 ('cross section*' OR questionnaire* OR survey OR surveys OR database or databases)):ti,ab,tt) NOT ('comparative study'/de OR 'controlled study'/de OR 'randomised controlled':ti,ab,tt OR 'randomized controlled':ti,ab,tt OR 'randomly assigned':ti,ab,tt)) OR ('cross‐sectional study'/de NOT ('randomized controlled trial'/de OR 'controlled clinical study'/de OR 'controlled study'/de OR 'randomised controlled':ti,ab,tt OR 'randomized controlled':ti,ab,tt OR 'control group':ti,ab,tt OR 'control groups':ti,ab,tt)) OR ('case control*':ti,ab,tt AND random*:ti,ab,tt NOT ('randomised controlled':ti,ab,tt OR 'randomized controlled':ti,ab,tt)) OR ('systematic review':ti,tt NOT (trial:ti,tt OR study:ti,tt)) OR (nonrandom*:ti,ab,tt NOT random*:ti,ab,tt) OR 'random field*':ti,ab,tt OR ('random cluster' NEAR/4 sampl*):ti,ab,tt OR (review:ab AND review:it) NOT trial:ti,tt OR ('we searched':ab AND (review:ti,tt OR review:it)) OR 'update review':ab OR (databases NEAR/5 searched):ab OR ((rat:ti,tt OR rats:ti,tt OR mouse:ti,tt OR mice:ti,tt OR swine:ti,tt OR porcine:ti,tt OR murine:ti,tt OR sheep:ti,tt OR lambs:ti,tt OR pigs:ti,tt OR piglets:ti,tt OR rabbit:ti,tt OR rabbits:ti,tt OR cat:ti,tt OR cats:ti,tt OR dog:ti,tt OR dogs:ti,tt OR cattle:ti,tt OR bovine:ti,tt OR monkey:ti,tt OR monkeys:ti,tt OR trout:ti,tt OR marmoset*:ti,tt) AND 'animal experiment'/de) OR ('animal experiment'/de NOT ('human experiment'/de OR 'human'/de))) AND [ENGLISH]/lim NOT ('case report'/de OR (case-report):ti) NOT ('juvenile'/exp NOT 'adult'/exp) NOT ([animals]/lim NOT [humans]/lim) NOT ([Conference Abstract]/lim OR [Conference Review]/lim)

**Medline**

(Non-alcoholic Fatty Liver Disease/ OR Fatty Liver/ OR (NAFLD OR MAFLD OR MASLD OR METALD OR FLD OR SLD OR MASH OR NASH OR ((nonalcoholic OR non-alcoholic OR metabol*) ADJ6 (fatty-liver* OR steatotic-liver* OR liver-steatos* OR hepat*-steatos* OR alcoholic-liver* OR liver-fibros* OR hepatic-fibros*)) OR ((fatty-liver* OR steato*) ADJ3 (diseas*)) OR Steatohepatit* OR ((fatty-liver* OR steatotic-liver* OR liver-steatos* OR hepat*-steatos* OR alcoholic-liver* OR liver-fibros* OR hepatic-fibros*) AND (hepatocellular-carcinom* OR HCC))).ab,ti,kf. OR ((metabol*) AND (fatty-liver* OR steatotic-liver* OR hepatic-steatos* OR alcoholic-liver* OR liver-fibros* OR hepatic-fibros* OR Steatohepatit*)).ti.) **AND** (exp Alcohol Drinking/ OR Alcoholism/ OR Drinking Behavior/ OR (((alcohol) ADJ3 (consumpt* OR intake OR "use" OR abuse OR drink*)) OR ((drink*) ADJ3 (behav* OR binge OR heavy))).ab,ti,kf.) **AND** (Exp Controlled clinical trial/ OR "Double-Blind Method"/ OR "Single-Blind Method"/ OR "Random Allocation"/ OR (random* OR factorial* OR crossover* OR cross over* OR placebo* OR ((doubl* OR singl*) ADJ blind*) OR assign* OR allocat* OR volunteer* OR trial).ab,ti,kf.) AND english.la. NOT (Case Reports/ OR (case-report).ti.) NOT ((exp Child/ OR exp Infant/) NOT (exp Adult/ OR exp Adolescent/)) NOT (exp animals/ NOT humans/) NOT (congres* OR abstract*).pt.

**Cochrane 443**

((NAFLD OR MAFLD OR MASLD OR METALD OR FLD OR SLD OR MASH OR NASH OR ((nonalcoholic OR non NEXT alcoholic OR metabol*) NEAR/6 (fatty NEXT liver* OR steatotic NEXT liver* OR liver NEXT steatos* OR hepat* NEXT steatos* OR alcoholic NEXT liver* OR liver NEXT fibros* OR hepatic NEXT fibros*)) OR ((fatty NEXT liver* OR steato*) NEAR/3 (diseas*)) OR Steatohepatit* OR ((fatty NEXT liver* OR steatotic NEXT liver* OR liver NEXT steatos* OR hepat* NEXT steatos* OR alcoholic NEXT liver* OR liver NEXT fibros* OR hepatic NEXT fibros*) AND (hepatocellular NEXT carcinom* OR HCC))):ab,ti,kw OR ((metabol*) AND (fatty NEXT liver* OR steatotic NEXT liver* OR hepatic NEXT steatos* OR alcoholic NEXT liver* OR liver NEXT fibros* OR hepatic NEXT fibros* OR Steatohepatit*)):ti) **AND** ((((alcohol) NEAR/3 (consumpt* OR intake OR use OR abuse OR drink*)) OR ((drink*) NEAR/3 (behav* OR binge OR heavy))):ab,ti,kw) NOT ("conference abstract":kw)

**Web of Science 517**

(TS=(NAFLD OR MAFLD OR MASLD OR METALD OR FLD OR SLD OR MASH OR NASH OR ((nonalcoholic OR non-alcoholic OR metabol*) NEAR/5 (fatty-liver* OR steatotic-liver* OR liver-steatos* OR hepat*-steatos* OR alcoholic-liver* OR liver-fibros* OR hepatic-fibros*)) OR ((fatty-liver* OR steato*) NEAR/2 (diseas*)) OR Steatohepatit* OR ((fatty-liver* OR steatotic-liver* OR liver-steatos* OR hepat*-steatos* OR alcoholic-liver* OR liver-fibros* OR hepatic-fibros*) AND (hepatocellular-carcinom* OR HCC)))) **AND** (TS=(((alcohol) NEAR/2 (consumpt* OR intake OR use OR abuse OR drink*)) OR ((drink*) NEAR/2 (behav* OR binge OR heavy)))) **AND** (TS=(random* OR factorial* OR crossover* OR (cross NEAR/1 over*) OR placebo* OR ((doubl* OR singl*) NEAR/1 blind*) OR assign* OR allocat* OR volunteer* OR trial)) NOT TS=((animal* OR rat OR rats OR mouse OR mice OR murine OR dog OR dogs OR canine OR cat OR cats OR feline OR rabbit OR cow OR cows OR bovine OR rodent* OR sheep OR ovine OR pig OR swine OR porcine OR veterinar* OR chick* OR zebrafish* OR baboon* OR nonhuman* OR primate* OR cattle* OR goose OR geese OR duck OR macaque* OR avian* OR bird* OR fish*) NOT (human* OR patient* OR women OR woman OR men OR man)) NOT TS=((juvenil* OR adolescen* OR preadolescen* OR youth* OR child* OR schoolchild* OR minors OR teen OR teens OR teenager* OR infan* OR toddler* OR pediatr* OR paediatr* OR puber* OR baby OR babies OR girl* OR boy* OR newborn* OR neonat* OR premature* OR pre-matur* OR kid OR kids OR underag* OR kindergar* OR pubescen* OR prepubesc* OR school* OR preschool* OR highschool* OR suckling OR PICU OR NICU OR PICUs OR NICUs) NOT (adult* OR elderl* OR man OR men OR woman OR women OR frail* OR octagener* OR geriat* OR ((old*) NEAR/3 (patient* OR peopl*)))) NOT DT=(Meeting Abstract OR Meeting Summary) AND LA=(English) NOT TI=(case-report)

**Google Scholar**

'nonalcoholic|metabolic fatty|steatotic liver'|'nonalcoholic liver|hepatic steatosis|fibrosis'|steatohepatitis 'alcohol consumption|intake|use|abuse' trial|RCT -animal -mouse -rat -child -children -pediatric -paediatric
